# CNV-ClinViewer: Enhancing the clinical interpretation of large copy-number variants online

**DOI:** 10.1101/2022.03.23.22272818

**Authors:** Marie Macnee, Eduardo Pérez-Palma, Tobias Brünger, Chiara Klöckner, Konrad Platzer, Arthur Stefanski, Ludovica Montanucci, Allan Bayat, Maximilian Radtke, Ryan L Collins, Michael Talkowski, Daniel Blankenberg, Rikke S Møller, Johannes R Lemke, Michael Nothnagel, Patrick May, Dennis Lal

## Abstract

**Purpose:** Large copy number variants (CNVs) can cause a heterogeneous spectrum of rare and severe disorders. However, most CNVs are benign and are part of natural variation in human genomes. CNV pathogenicity classification, genotype-phenotype analyses, and therapeutic target identification are challenging and time-consuming tasks that require the integration and analysis of information from multiple scattered sources by experts.

**Methods:** We developed a web-application combining >250,000 patient and population CNVs together with a large set of biomedical annotations and provide tools for CNV classification based on ACMG/ClinGen guidelines and gene-set enrichment analyses.

**Results:** Here, we introduce the CNV-ClinViewer (https://cnv-ClinViewer.broadinstitute.org), an open-source web-application for clinical evaluation and visual exploration of CNVs. The application enables real-time interactive exploration of large CNV datasets in a user-friendly designed interface.

**Conclusion:** Overall, this resource facilitates semi-automated clinical CNV interpretation and genomic loci exploration and, in combination with clinical judgment, enables clinicians and researchers to formulate novel hypotheses and guide their decision-making process. Subsequently, the CNV-ClinViewer enhances for clinical investigators patient care and for basic scientists translational genomic research.

## INTRODUCTION

Copy-number variation (CNV) disorders arise from the dosage imbalance of one or more genes, resulting from deletions, duplications, or other genomic rearrangements. Most disease-associated CNVs are unique, cause severe complex disorders and are often hard to distinguish from benign CNVs frequently found in the general population^1^.

With an average size of pathogenic/likely pathogenic CNVs in ClinVar^2^ of 9.7 Mb (95% CI 9.3-10.1 Mb, median=2.6 Mb), disease-associated CNVs typically cover tens to hundreds of genes. Thus, pinpointing the underlying disease driver and modifier genes represents a major challenge. Given the complexity of CNV interpretation, the American College of Medical Genetics and Genomics (ACMG) has developed technical standards (a quantitative, evidence-based scoring framework) to standardize the evaluation process of the genomic content of a CNV region, and to promote consistency and transparency in classification and reporting across clinical laboratories^1^. The application of those standards involves clinical and genetic CNV data that are scattered across registries, databases, and the literature. Therefore, it is difficult and highly time-consuming to annotate, analyze and interpret CNVs manually in a clinical setting.

Several bioinformatics tools, often fully-automated, have been developed for clinicians and researchers to facilitate large-scale CNV classification according to the ACMG guidelines (for example see^3,4^). However, no algorithm or even classification framework can perfectly capture data that is usually only available to experts, such as deep clinical, CNV, or gene-level information. This specifically affects data that cannot be extracted from public online resources, such as detailed information about the phenotype, including family history, CNV inheritance pattern, and many other criteria currently not integrated in the clinical significance classification. For the semi-automated tools^5^, the extracted and manually entered information is not explorable within the same web-application interface and comparative visual inspection of CNVs with various genomic and clinical data sources is not possible because current tools do not provide such functionality^3–5^.

While the current focus of those existing tools is on clinical significance classification, they are not designed to perform ad-hoc evaluations of new data such as genotype-phenotype analyses to expose undiscovered patterns of CNV localization in patient cohorts. To show a correlation of patient CNVs with known disease genes at a specific locus or to narrow down dosage sensitive genes that are likely driver genes in particular CNVs, the interactive visual inspection of affected genomic content combined with crosslinked data from various sources can enhance the interpretational analysis of CNVs. Paradoxically, the individuals who collect the most relevant clinical information for CNV clinical significance classification and interpretation, such as genetic counselors, treating physicians, and patients’ families, often lack the required bioinformatical expertise to examine patient CNVs thoroughly. To overcome current limitations for biomedical CNV interpretation of those individuals and researchers, we developed the CNV-ClinViewer, a fully open-source exploration and interpretation platform that integrates analysis, annotation, guideline-based, classification and clinical evaluation of CNVs and provides users, even families with CNVs of uncertain significance, with an interface to curate, visualize, interact with and continuously re-evaluate the CNV data.

## MATERIAL AND METHODS

Details about the data sources used in the CNV-ClinViewer can be found in the **Supplementary Table 1**.

### Genetic CNV variants and regions datasets

Patient CNVs were obtained from ClinVar^2^ (updated quarterly), and CNVs identified in the general population were obtained from the UK biobank^6^ as well as from gnomAD^7^ (controls-only dataset, version 2.1). GnomAD CNVs were reduced to those with a filter equal to PASS, and SVTYPE equivalent to ‘DEL’ or ‘DUP’, and all CNVs from the general population were filtered for large CNVs >50kb. Specifically, we collected 52,344 CNVs from ClinVar (10654 pathogenic/ likely pathogenic CNVs), 195,916 CNVs from the UK-biobank (>50kb), and 10,370 CNVs from gnomAD (>50kb).

To annotate neutral as well as clinically relevant regions by use of a visual summary of the variants, we further processed the variants by counting the number of deletions or duplications in the different data sets using a sliding window approach with a window size of 200kb and step size of 100kb. For the cohort-level data from the UK-biobank and gnomAD, we divided the counts by the number of samples in the cohort to retrieve region-specific allele-frequencies.

Known CNV syndromes and genomic regions with evaluated dosage sensitivity were obtained from DECIPHER^8^ and ClinGen^9^ (updated quarterly) databases.

### Gene-level annotations

We retrieved a list of 18,792 protein-coding gene symbols from the HUGO Gene Nomenclature Committee (HGNC)^10^. These genes were annotated by the genomic boundaries of one transcript from RefSeq^11^ (hg19), either the MANE select transcript or alternatively the longest transcript. In addition, we annotated gene-level scores for sequence constraint and dosage sensitivity in humans (**Table 1**) and gene-disease associations from ClinGen^9^ (updated quarterly).

**Table 1.**
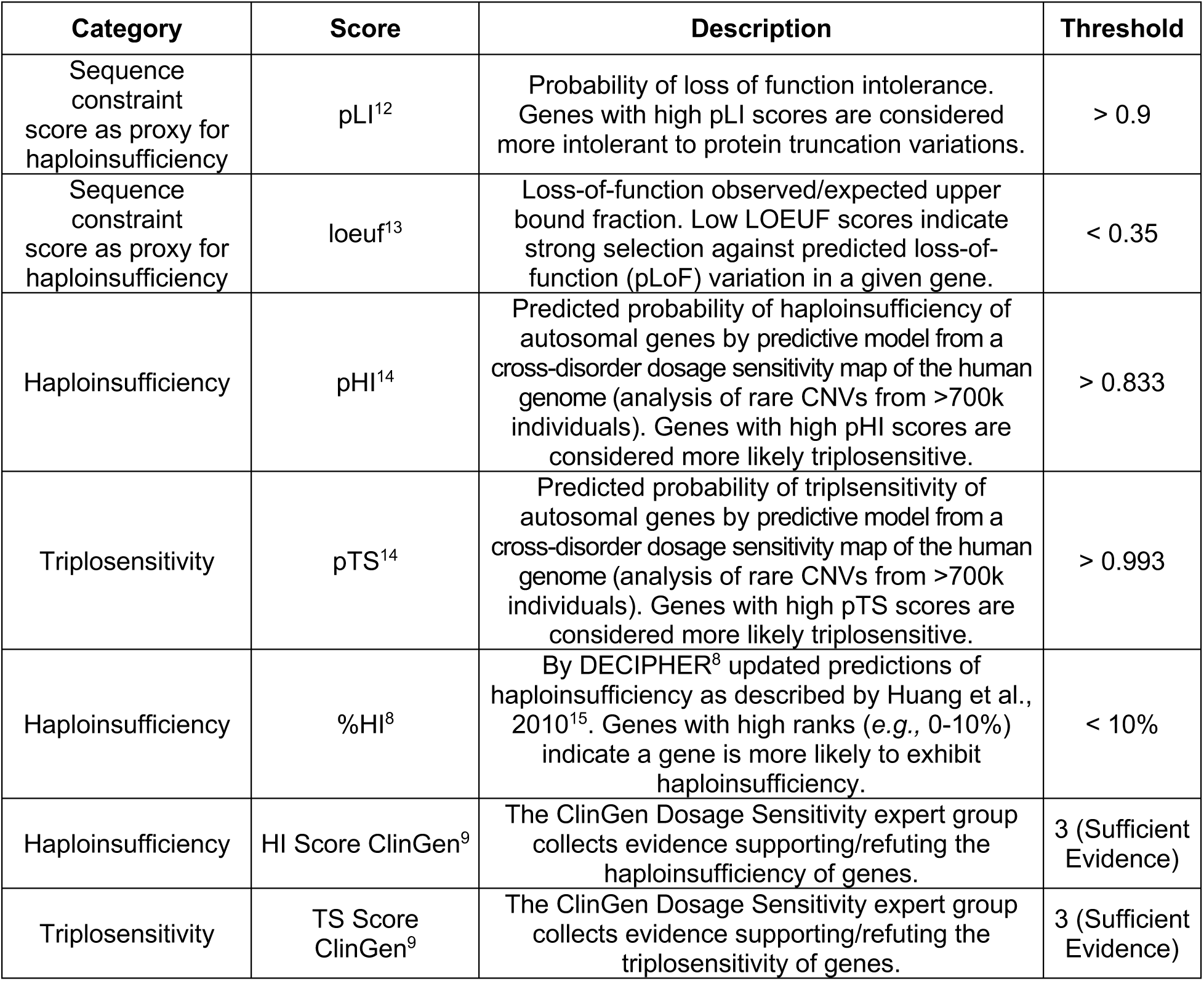
Description of gene-level scores for sequence constraint and dosage sensitivity. The given thresholds are used in the CNV-ClinViewer to annotate dosage sensitivity of genes.

### CNV-ClinViewer web server development

The CNV-ClinViewer was developed with the Shiny framework of R studio software (v.1.7.1, https://shiny.rstudio.com/) which transforms regular R code into an interactive environment that can follow and ‘react’ to remote-user instructions. The pre-processed data alongside the R/Shiny code was uploaded as a stand-alone Ubuntu image with Google Cloud services. The image was deployed into a Google Virtual Machine (VM) using the googleComputeEngineR package (v.0.3.0, https://github.com/cloudyr/googleComputeEngineR). The CNV-ClinViewer web server (https://cnv-ClinViewer.broadinstitute.org/) is compatible with all commonly-used internet browsers.

For the classification of uploaded CNVs, the clinical interpretation tool ClassifyCNV^3^ including its own data sources was integrated into the CNV-ClinViewer framework. Upon selection of one of the uploaded and classified CNVs, all pre-processed data are intersected by genomic coordinates of the selected CNV. Consequently, the HTML report describing the selected CNV is generated using the rmarkdown R package (v2.11, https://rmarkdown.rstudio.com), and the interactive visualizations and tables are rendered using the ggplot2 (v.3.3.5, https://ggplot2.tidyverse.org), plotly (v.4.9.4.1 https://plotly-r.com) and DT (v.0.19, https://CRAN.R-project.org/package=DT) R packages. For the gene set enrichment analysis, the enrichr R package (v3.0, https://CRAN.R-project.org/package=enrichR) that provides an interface to the Enrichr database is used^16^.

The CNV-ClinViewer is an open-source project, and its code will continue to grow and improve through version control in the GitHub repository (https://github.com/LalResearchGroup/CNV-clinviewer).

## RESULTS

We present the CNV-ClinViewer, a user-friendly web-application that semi-automatedly enhances the ACMG guideline-based CNV classification for caregivers and facilitates the biomedical interpretation of gene content at a locus of interest for scientists. Development, workflow, and main features are presented in **Figure 1**.

**Figure 1.**
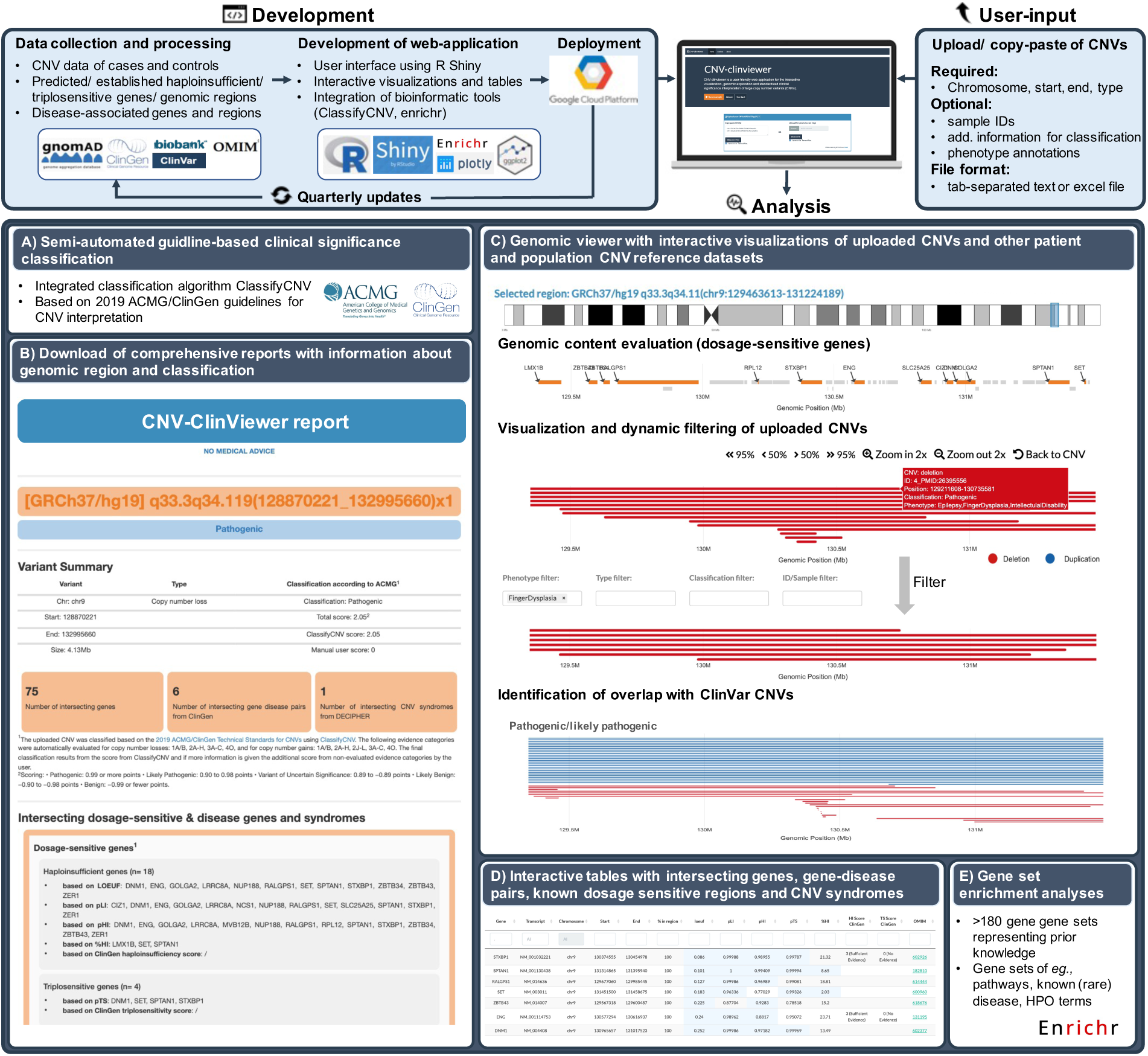
CNV-ClinViewer overview. CNV data as well as clinical and genomic annotations are collected, processed and annotated, and quarterly updated. Users can copy-paste CNV(s) or upload CNVs in a tab-separated or Excel file for real-time exploration and interpretation. The key features of the CNV-ClinViewer are depicted in A-E. **A)** The integrated *semi-automated classification* of uploaded CNVs is based on 2019 ACMG/ClinGen Technical Standards for CNVs by ClassifyCNV^3^ and is one of the key features of the CNV-ClinViewer. The resulting scores and applied evidence categories can be inspected in a table overview and also downloaded. **B)** A *comprehensive report on individual CNVs*, including details about the clinical significance classification and the overlap with established/predicted haploinsufficient/triplosensitive and clinically relevant genes and genomic regions, can be downloaded. **C)** A *genomic viewer* allows inspecting the uploaded CNVs and their genomic region alongside biomedical annotations and other pathogenic and general population CNV datasets. Here, the user can visually compare and dynamically filter uploaded CNVs, perform a seamless evaluation of the gene content, identify disease-prone regions and find unknown patterns of CNV localization to generate hypotheses for further research. An extended view of the genomic viewer can be found in Supplementary Figure 1. **D)** *Dynamic interaction*: users can retrieve, filter, and download information about the intersecting genes, gene-disease associations, known dosage sensitive regions, and known CNV syndromes. **E)** Users can perform *gene-set enrichment analyses* (GSEA) to infer information on genes within a selected genomic region or CNV by comparing it to >180 annotated gene sets^16^ representing prior biological knowledge such as pathways, Human Phenotype Ontology (HPO) terms and known (rare) diseases.

### Upload of one up to thousands of CNVs for clinical interpretation and genetic reports

The CNV-ClinViewer allows simultaneous analysis of single or multiple CNVs, irrespective of the technology used to identify them. It requires as input the genomic coordinates of CNVs based on the human reference genome GRCh37/hg19 (https://www.ncbi.nlm.nih.gov/assembly/GCF_000001405.13/), either by copy-pasting or by uploading a tab-separated text or Excel file (max. number of CNVs = 10,000). Minimal required information for each CNV, including whole chromosome trisomies and monosomies, is the chromosome, start, end and CNV type (deletion or duplication). Optionally, the user can provide sample IDs, phenotype information for filtering and a score of manually assessed evidence categories of the 2019 ACMG standard guidelines (see below). Details and example files can be found in the help section and on the about page of the CNV-ClinViewer.

After CNV submission, the user is directed to the analysis interface. Here, five different analysis panels are available. In the *first analysis panel*, the uploaded CNVs are displayed in a downloadable table comprising their annotated clinical significance and details about the scoring (**Figure 1A**). The CNVs are classified by ClassifyCNV^3^, a command-line tool integrated into the CNV-ClinViewer infrastructure that automatically evaluates the evidence categories 1A/B, 2A-H, 3A-C, 4O for copy-number losses and 1A/B, 2A-H, 2J-L, 3A-C, 4O for copy-number gains from the 2019 ACMG/ClinGen Technical Standards for CNVs^1^. In case the user uploaded additional scores of relevant ACMG clinical significance criteria for which additional information, such as family history or the *de novo* status, is required, the score is added to the score from ClassifyCNV to refine the final score and classification. In the *second analysis panel*, a comprehensive report of the individual CNVs can be downloaded (**Figure 1B**). The report includes details about the clinical significance classification and the overlap with established/predicted haploinsufficient/triplosensitive and clinically relevant genes and genomic regions.

### Uploaded CNV data can be visually inspected alongside publicly available data

The *third panel* of the analysis interface is a genomic viewer (**Figure 1C, Supplementary Figure 1A-F**). Here, the uploaded CNVs and their genomic region can be inspected alongside biomedical annotations and other pathogenic and general population CNV datasets. Included are five data tracks that the user can interactively navigate by zooming in/out, moving horizontically, and selecting genomic regions of interest. In the first track of the genomic viewer (**Supplementary Figure 1B**), the CNV-ClinViewer integrates seamless evaluation of the gene content by visualization of all protein-coding genes. Here, the genes are highlighted by a selection of gene dosage sensitivity scores (**Table 1**) to enable fast and visual gene prioritization in context to the uploaded CNVs. Below this gene track, all uploaded CNVs that intersect the selected region are visualized and can be interactively filtered based on uploaded phenotypic annotations, their assigned clinical significance, CNV type, and sample IDs (**Supplementary Figure 1C**). The further tracks enable the visual identification of disease-prone regions with coupled visualizations of summarized data (allele frequencies and allele counts) of >250,000 CNVs from the general population (UK Biobank^6^, gnomAD^7^) and pathogenic/likely pathogenic CNVs from ClinVar^2^ (**Supplementary Figure 1D-F**). In addition, users can identify the overlap of uploaded CNVs with individual ClinVar CNVs (**Supplementary Figure 1D**). The latter are displayed in an interactive plot grouped by their assigned clinical significance (pathogenic/likely pathogenic vs. uncertain significance vs. benign/likely benign) and can be filtered based on their type and clinical significance. Details about the ClinVar CNVs, such as their reported phenotypes or allele origin and the link to the ClinVar variant website, are shown in a table and can be downloaded for further analyses.

### Additional features and analyses provide users with advanced CNV insights

Below the genomic viewer, the user can retrieve and download information about the intersecting genes, classified gene-disease associations from ClinGen^9^, known dosage sensitive regions and CNV syndromes (**Figure 1D**). The gene table contains all protein-coding genes, their genomic coordinates (hg19/GRCh37) and transcript IDs, links to the Online Mendelian Inheritance in Man database (OMIM, https://omim.org/), and gene dosage sensitivity scores (**Table 1**). All genes classified by ClinGen with reported associations with one or several diseases are shown in the gene-disease association table, including links to the comprehensive gene-disease association reports on the ClinGen website. In the ClinGen region table and the DECIPHER^8^ CNV syndrome table, all well-characterized dosage sensitive regions and developmental CNV syndromes that intersect a CNV/genomic region of interest can be retrieved.

In the last panel, the user can perform gene set enrichment analyses to infer knowledge about the genes from a selected genomic region by comparing it to >180 annotated gene sets representing prior biological knowledge such as pathways, Human Phenotype Ontology (HPO) terms^17^, and known (rare) diseases (**Figure 1E**).

### Example analysis: 9q33.3q34.11 microdeletions

To illustrate the utility of the CNV-ClinViewer, we performed an example analysis of 14 previously published CNVs (9q33.3q34.11 microdeletions) from patients with complex developmental disorders from the literature (n=14)^18–21^. Here, the CNVs were first uploaded and automatically classified as (likely) pathogenic (**Supplementary File 1: Figure 1-2**). Next, the smallest region of overlap with its dosage sensitive gene *STXBP1* could be immediately identified due to the interactive visualization of the overlapping patient CNVs (**Supplementary File 1: Figure 3-4**). In addition, the CNV-ClinViewer could resolve the clinical heterogeneity of patients by enabling the user to filter the CNVs based on assigned phenotypes (**Supplementary File 1: Figure 6**) and retrieve region-specific information about intersecting clinically relevant genes and their associated phenotypes (**Supplementary File 1: Figure 5**) as well as overlapping patient CNVs from ClinVar (**Supplementary File 1: Figure 7**). Overall, we demonstrate the CNV-ClinViewer’s fast and seamless biomedical interpretation of CNVs and its potential to replicate research findings due to a large number of annotations and visual inspection capabilities. To perform a comparable analysis without the CNV-ClinViewer, the user would be required to classify the CNVs manually one at a time or with the support of an existing tool, visualize the CNVs with yet another tool or genomic viewer, visit several webpages, and process and annotate information from various databases. The example analysis can be found in more detail and with step-by-step instructions illustrated with images in the **Supplementary File 1**.

## DISCUSSION

By aggregating >250,000 population and patient CNVs, annotating various gene-scores and clinical annotations, and integrating useful existing bioinformatics tools, we developed a novel and user-friendly interface as a decision support tool for the clinical evaluation of CNVs and comparative visual inspection.

CNV analysis and interpretation tools and, in general, clinical decision support tools can make patient care more efficient, cost-effective, and guideline-concordant. However, although the prediction results from many approaches are promising, their value is limited by their lack of interpretability and human intuition. The black-box nature of some pathogenicity predictions can further exacerbate trust issues and worsen the overall experience^22^. To address this issue, human-centered tools with interactive mechanisms can grant end-users more agency in guiding the interpretation and can be used for critical decision-making purposes beyond an algorithm^22,23^. Therefore, in the development of the CNV-ClinViewer, we emphasized interactive analyses of the CNVs beyond a classification score and allow the user to dynamically inspect and compare the results in detail. In comparison, existing open-source tools^3–5^ focus more on the classification scores than on the interactive inspection and further investigations. We believe that the comprehensive approach of the CNV-ClinViewer will empower users with more trust in the results, the agency to test hypotheses, and enable them to apply their domain knowledge while simultaneously leveraging the benefits of automation.

To ensure that a tool is easy to use and is designed for target users at each development stage, human-centered design methods and principles should be applied^23,24^. Such methods draw from well-established design principles in many disciplines, including usability, visualizations, and interface design, and consider users’ context and experience. As the effectiveness of clinical tools may be severely limited without application of those principles and could even contribute to adverse medical events^25^, we put great emphasis on developing a user-friendly interface with simple data upload with minimal requirements, a clear analysis workflow, and real-time results, while giving the user the ability to retrieve details and help, and modify the analysis when required. We also focused on the visual and intuitive inspection of CNV data and its interaction with the user. In particular, for genomic data, the integration of visualizations is essential for interpretation and hypothesis generation, essential for combining different data sources and discovering unexpected localization patterns of genomic annotations in a short amount of time, as well as a valuable aid in communicating discoveries^26^.

On the one hand, the CNV-ClinViewer will assist clinicians, clinical geneticists and genetic counselors analyzing and interpreting CNV data and improve objectivity and accuracy in the clinical work-up. On the other hand, the collected and integrated knowledge combined with clinical judgment will enable clinicians and researchers to formulate novel hypotheses and guide their decision-making process.

The current features of the tool are focused on large CNVs that intersect several genes when gene prioritization plays a role in the analysis. Future versions will increase the granularity of the tool, for example, by providing a more detailed gene track that includes exons and introns, and different transcripts with expression data. With that, it will bring more value for interpreting intragenic deletions and duplications. Also, to integrate with existing workflows, we plan to develop an application programming interface (API) to query the system and fetch the results without using a graphical user interface. Overall, the CNV-ClinViewer is the first framework that enables interactive exploration of CNV data and semi-automated variant classification. With its scalable architecture, the web-application is well suited for both genome-wide (re-)evaluations of large data sets and small number of CNVs. The CNV-ClinViewer is an open-source tool (https://www.cnv-ClinViewer.broadinstitute.org/), including complete documentation on the use of the website.

## Data Availability

All code and data are accessible at

https://github.com/LalReserachGroup/CNV-clinviewer

## DATA AVAILABILITY

The data used in the CNV-ClinViewer are listed in Supplementary Table 1. All code and data are accessible at https://github.com/LalReserachGroup/CNV-clinviewer, and detailed documentation can be found at https://www.cnv-ClinViewer.broadinstitute.org/. The web-application is free and open to all users, and there is no login requirement.

## ACKNOWLEDGEMENTS

**E.P**. is supported by Agencia Nacional de Investigación y Desarrollo (ANID, PAI77200124) of Chile and the FamilieSCN2A foundation 2020 Action Potential Grant.

## AUTHOR CONTRIBUTIONS

Conceptualization: D.L., M.M., E.P.; Data curation: M.M., E.P.; Methodology: D.L., M.M., E.P., P.M., M.N.,; Supervision: D.L., E.P.; Validation: T.B., C.K, K.P., A.S., L.M., A.B., M.R., R.C., D.B., J.L., M.T., R.S.M.; Visualization: M.M.; Writing-original draft: D.L., M.M., E.P.; Writing-review & editing: P.M., M.N., E.P., T.B., C.K, K.P., A.S., L.M., A.B., M.R., R.C., D.B., J.L., M.T., R.S.M.

## CONFLICT OF INTEREST

Disclosure: The authors declare no conflict of interest.

## SUPPLEMENTARY DATA

**Supplementary Table 1.**
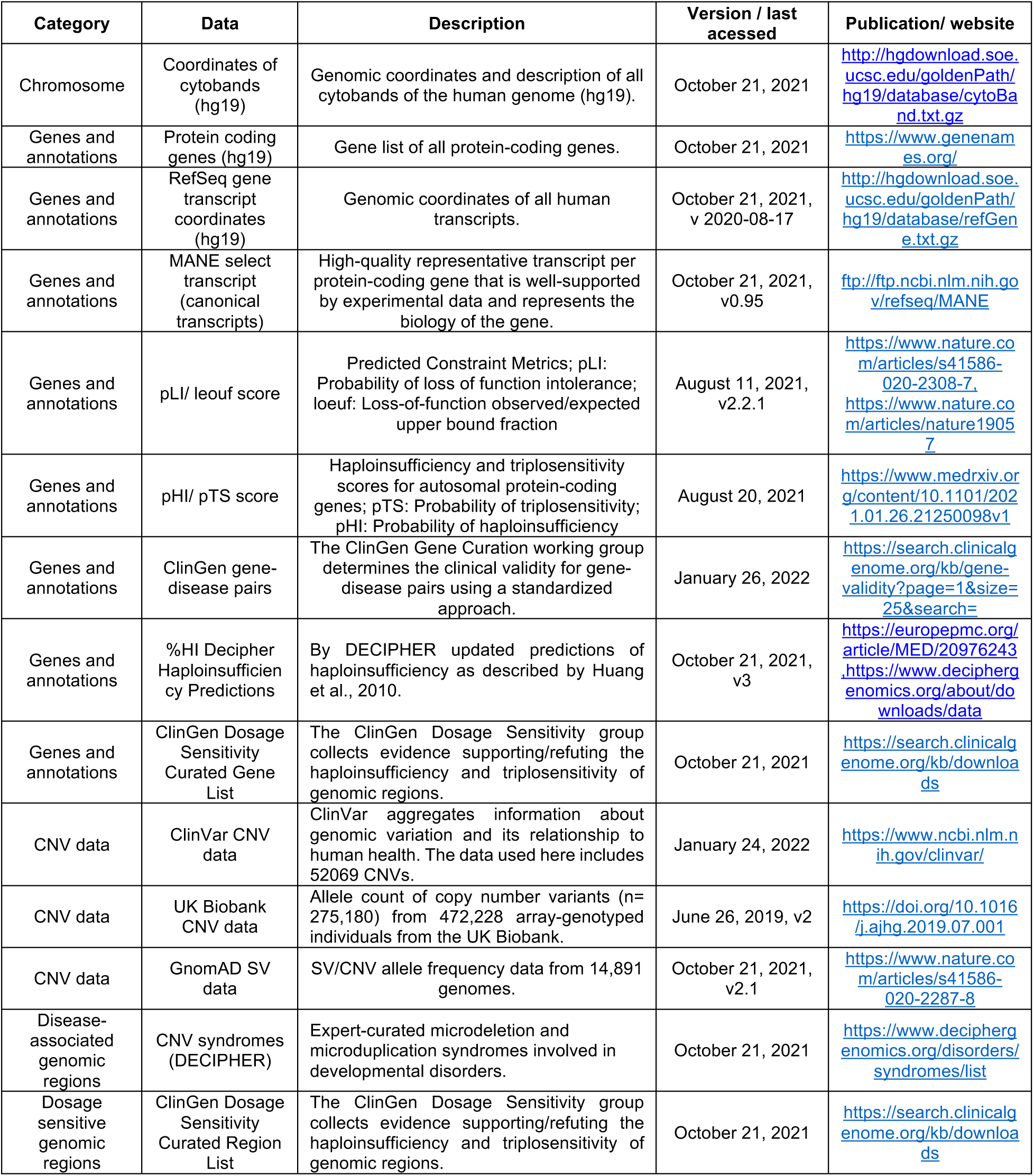
Data sources.

**Supplementary Figure 1.**
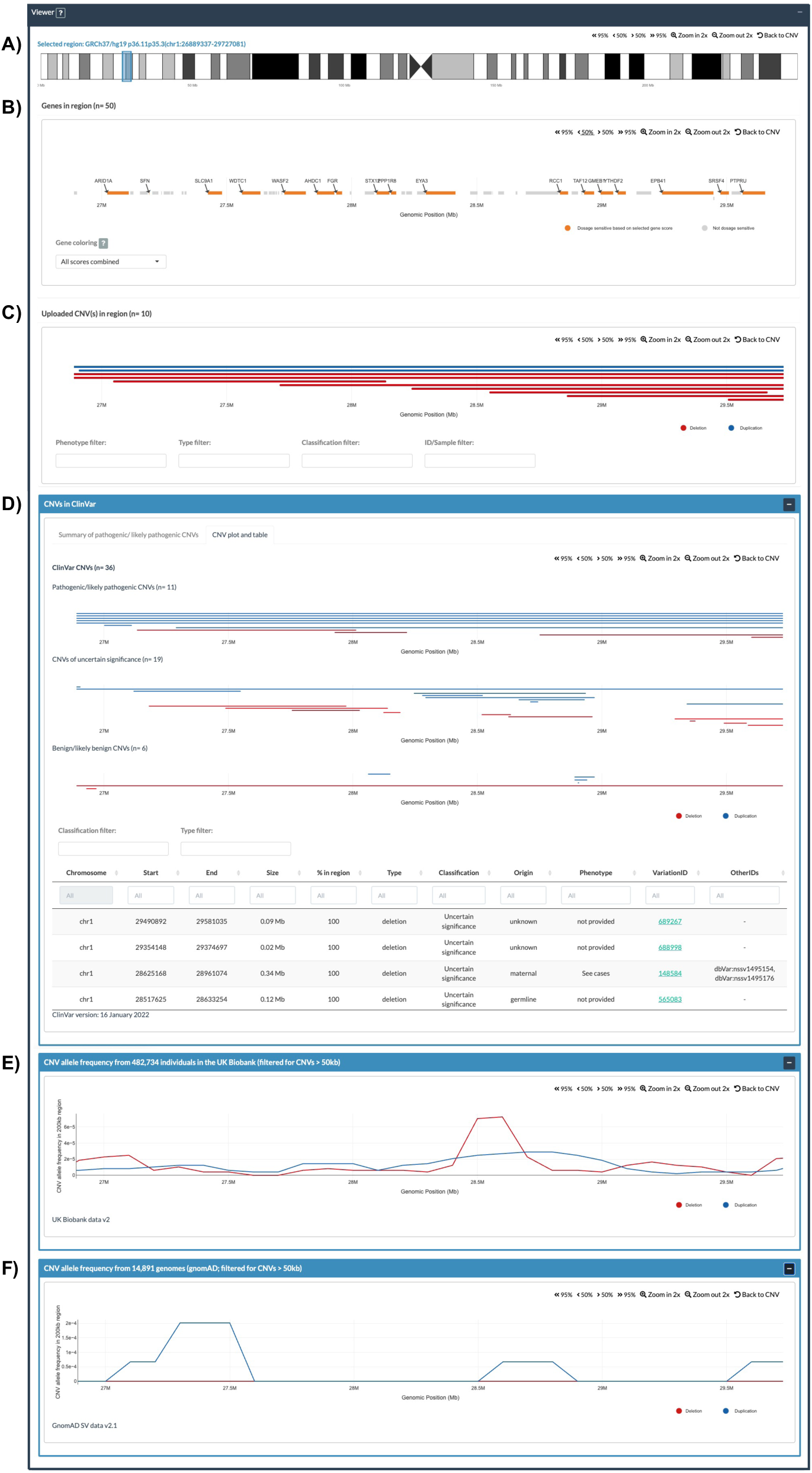
Genomic Viewer of CNV-Clinviewer. The genomic viewer enables the inspection of the uploaded CNVs and their genomic region alongside biomedical annotations and other pathogenic and general population CNV datasets. Users can interactively navigate by zooming in/out, moving, and selecting genomic regions of interest. **A)** The ideogram shows the selected genomic region. Users can select a genomic region of interest by Drag’n’Drop. **B)** In the gene track, all protein-coding genes are shown. Dosage-sensitive genes, based on a selection of different gene dosage sensitivity scores, are highlighted in orange for gene prioritization in context to the uploaded CNVs. **C)** Uploaded CNVs that intersect the selected region are visualized and can be interactively filtered based on uploaded phenotypic annotations, their assigned clinical significance, CNV type, and sample IDs. Deletions are visualized as red, duplications as blue bars. **D)** The ClinVar track has two tabs. The first tab (not shown) shows a visual summary of allele counts from pathogenic/ likely pathogenic CNVs. The second tab displays all CNVs from ClinVar in a plot grouped by their assigned clinical significance (pathogenic/ likely pathogenic vs. uncertain significance vs. benign/ likely benign) and can be filtered based on their type and clinical significance. Details about the ClinVar CNVs, such as their reported phenotypes or allele origin and the link to the ClinVar variant website, are shown in a table and can be downloaded for further analyses. **E)** Visualizations of summarized allele frequencies of large CNVs (>50kb) from the UK Biobank and **F)** gnomAD.

## Notes

### Competing Interest Statement

The authors have declared no competing interest.

### Funding Statement

This study did not receive any funding

### Author Declarations

The study used (or will use) ONLY openly available human data that were originally located at: - https://www.ncbi.nlm.nih.gov/clinvar/ (CNV data from ClinVar) -https://biobankenginedev.stanford.edu/downloads (CNV data from UK biobank) - https://gnomad.broadinstitute.org/downloads/ (gnomAD SV data)

