## Supplementary File 1 for "CNV-ClinViewer: Enhancing the clinical interpretation of large copy-number variants online"

To illustrate the utility of the CNV-ClinViewer with a proof-of-concept example, we used 9q33.3q34.11 microdeletions from the literature (n=14), along with the phenotypic status of phenotypes with high prevalence in the patient population (epilepsy, intellectual disability, finger dysplasia, facial dysmorphism)[1-4]. After uploading the data (Fig. 1), all CNVs are displayed in the table with their automatically assigned pathogenic (12/14 CNVs) or likely pathogenic (2/14 CNVs) classification (Fig. 2).

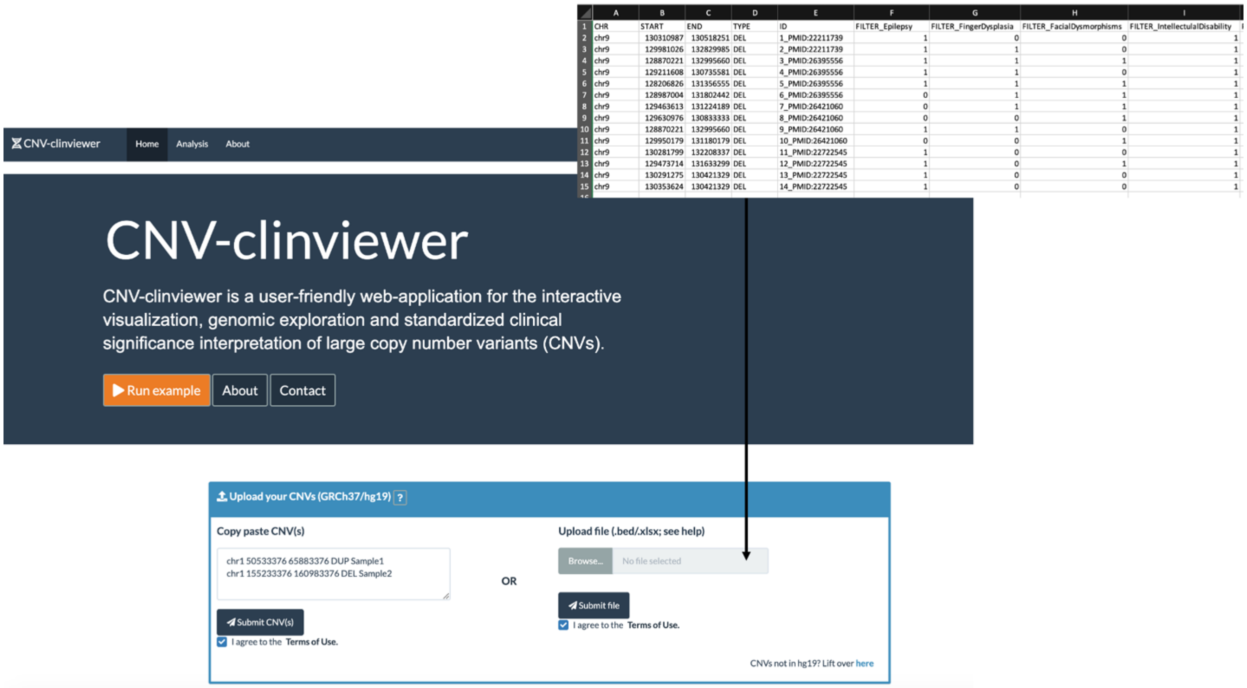


**Fig. 1: CNV data upload.** The data of 9q33.3q34.11 microdeletions (n=14) is uploaded as an Excel file with the required chromosome, start, end and type data, and additionally with sample IDs and binary phenotypes for filtering.

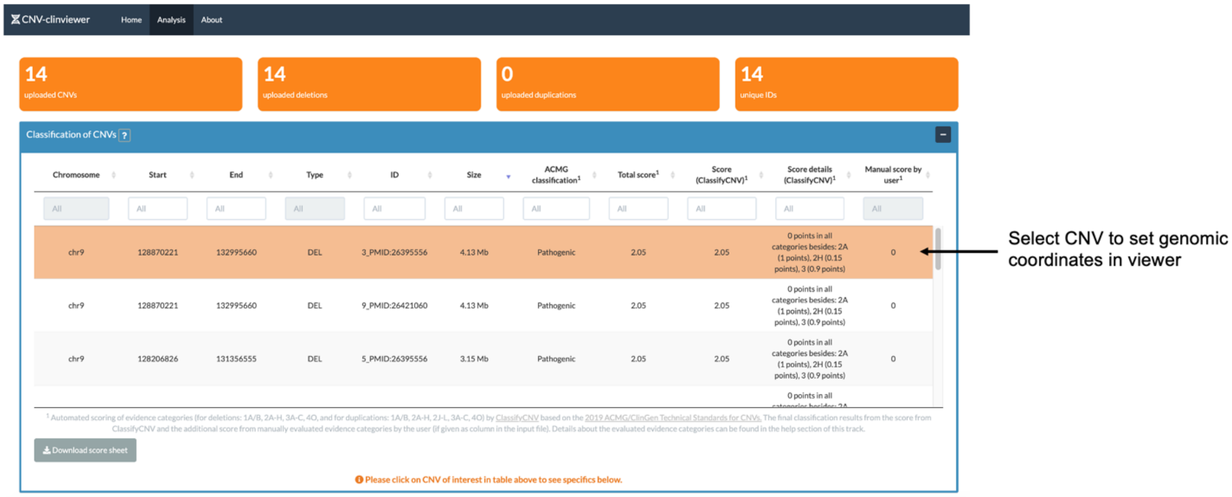

**Fig. 2: Table of uploaded and classified CNVs.** Upon upload of the data, the CNVs are classified by ClassifyCNV[5] based on the 2019 ACMG/ClinGen guidelines and displayed in a table, including details about the scoring. Further information about the scoring of the evidence categories by ClassifyCNV can be retrieved in the help section of the panel by pressing the button next to the title of the panel. CNVs of interest can be selected in the table to download a comprehensive report and explore its genomic region and content in the genomic viewer.

From this table, we selected the largest CNV to set the genomic coordinates of interest for the genomic viewer and inspected the visualization of the uploaded CNVs and intersecting genes (Fig. 3). Here, one can observe that all CNVs overlap while their size is variable and that 18 genes in this region (e.g., *STXBP1* and *SPTAN1*) indicate dosage sensitivity by one or more of the integrated dosage sensitivity scores (highlighted in orange in the gene visualization).

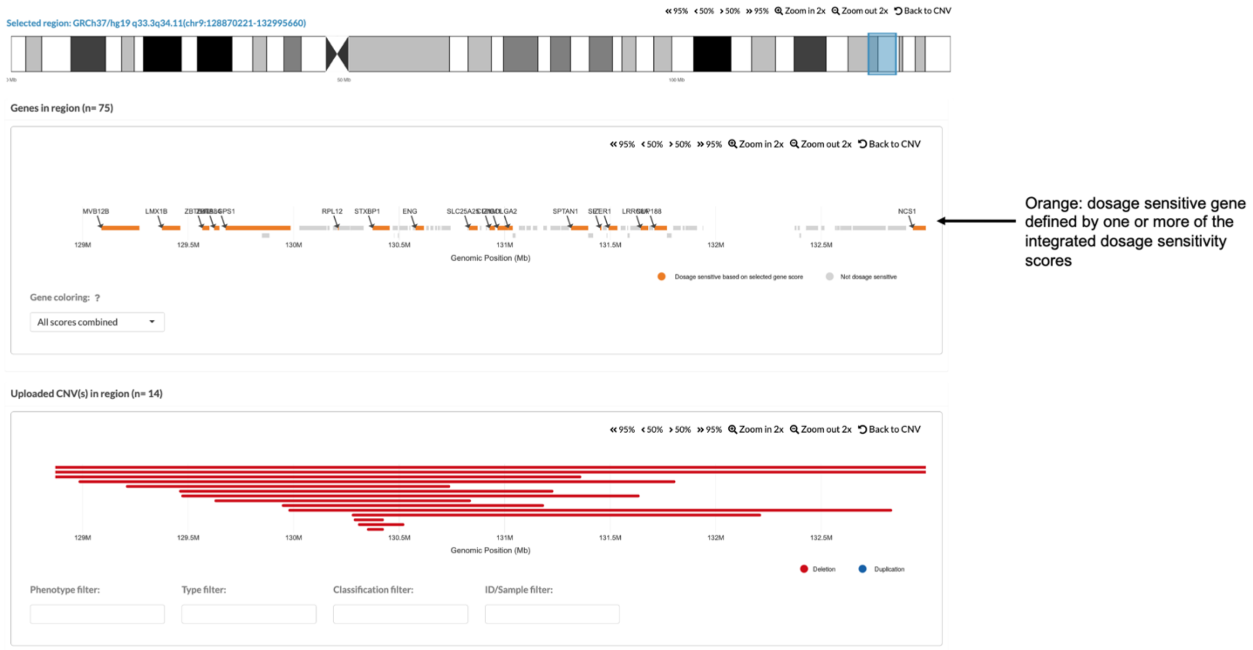


**Fig. 3: Genomic viewer with visualization of uploaded CNVs and intersecting genes.** Deletions are shown in red, and duplications are shown in blue (in this example there are only deletions). Dosage sensitive genes are displayed in orange to support the gene prioritization visually. The user can select from several dosage sensitive scores or can choose a combination of all.

To identify which gene(s) intersect the smallest region of overlap, we simply selected the plot area by Drag'n'Drop and identified the gene *STXBP1* (Fig. 4). By inspecting the individual dosage sensitivity scores in detail in the tooltip by hovering over the gene (Fig. 4) or in the gene table (Fig. 5), we found that all scores of *STXBP1* indicate dosage sensitivity, *e.g.*, the haploinsufficiency score of 3 (= sufficient evidence) from ClinGen.

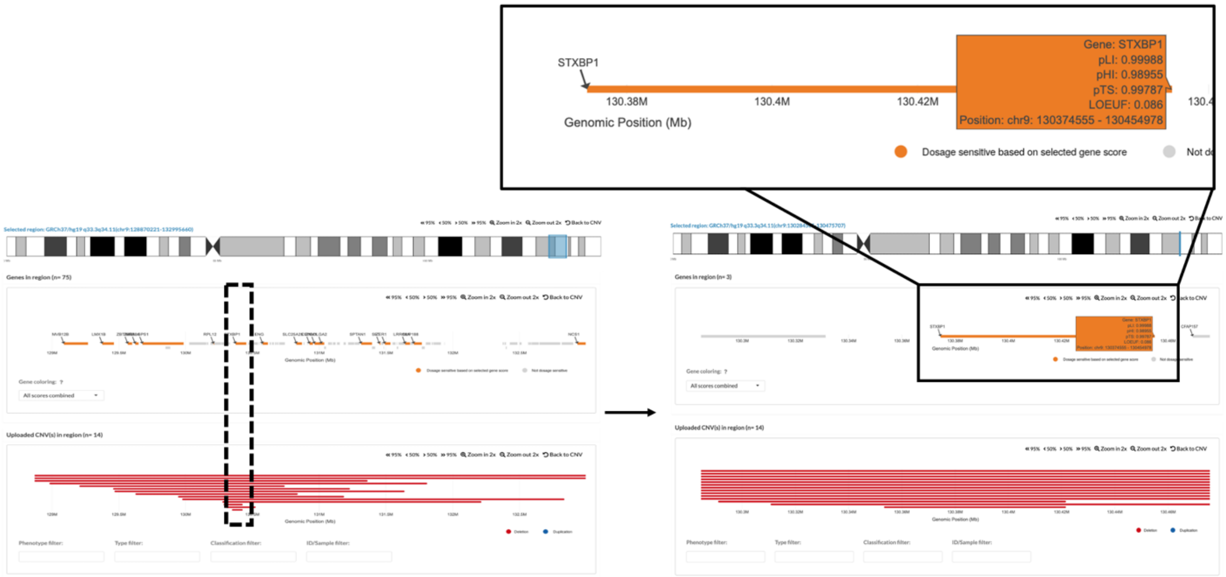


**Fig. 4: Identification of gene content in the smallest region of overlap of uploaded CNVs.** In order to zoom in the smallest region of overlap, the user can select the plot area by Drag'n'Drop. By hovering over genes and CNVs, the user can retrieve more information about those in a tooltip.

In addition, the table of intersecting gene-disease pairs from ClinGen lists *STXBP1* to be associated with developmental and epileptic encephalopathy (DEE) and links to the ClinGen online report (Fig. 5). Indeed, the gene is defined as one of the main causes of Ohtahara syndrome (OS), a devastating early onset DEE, and known as primary driver of 9q33.3q34.11 microdeletions[4].

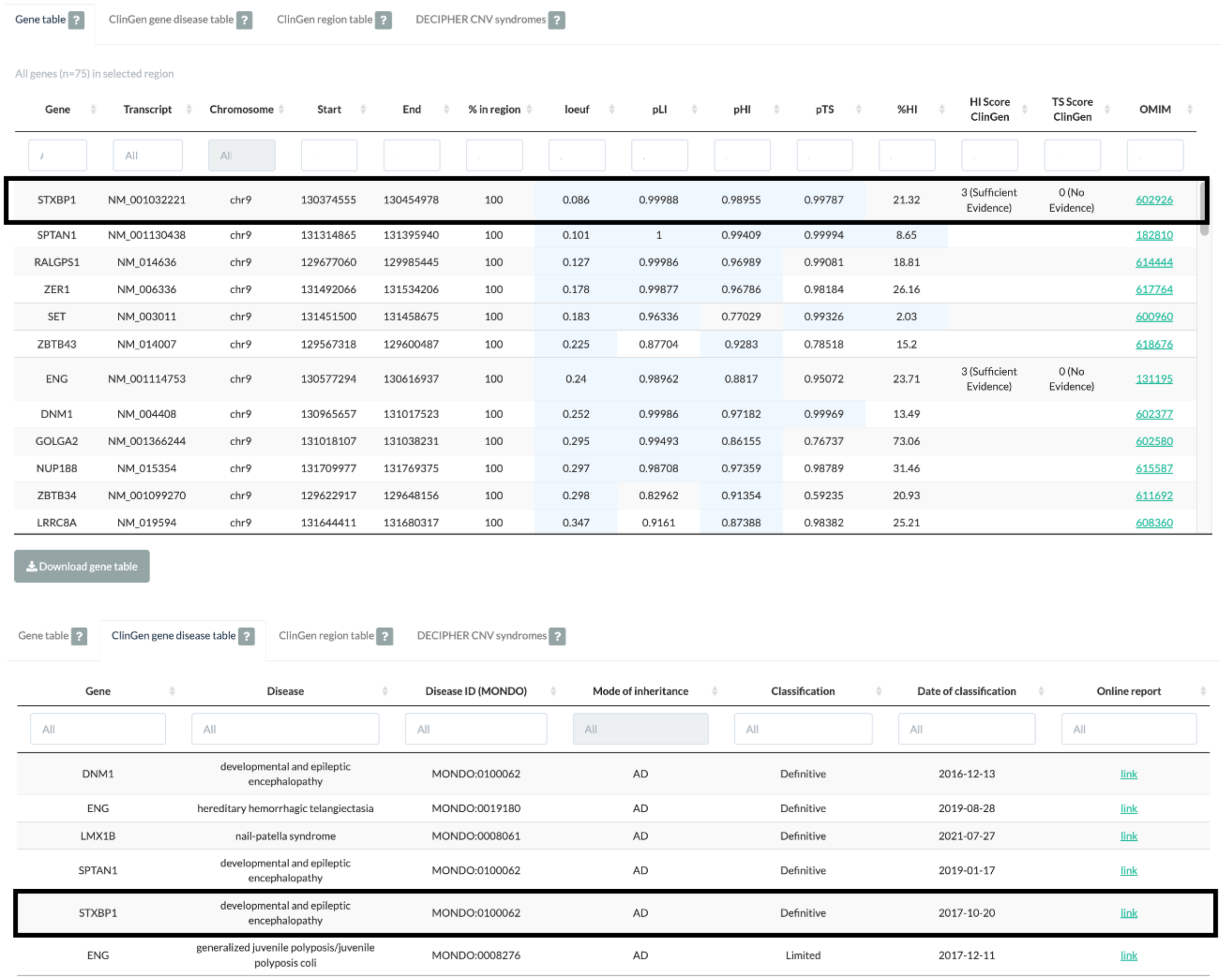
Gene table ? ClinGen gene disease table ? ClinGen region table ? DECIPHER CNV syndromes ?

All genes (n=75) in selected region

| Gene | Transcript | Chromosome | Start | End | % in region | loeuf | pLI | pHI | pTS | %HI | HI Score ClinGen | TS Score ClinGen | OMIM |
|---|---|---|---|---|---|---|---|---|---|---|---|---|---|
| STXBP1 | NM_001032221 | chr9 | 130374555 | 130454978 | 100 | 0.086 | 0.99988 | 0.98955 | 0.99787 | 21.32 | 3 (Sufficient Evidence) | 0 (No Evidence) | 602926 |
| SPTAN1 | NM_001130438 | chr9 | 131314865 | 131395940 | 100 | 0.101 | 1 | 0.99409 | 0.99994 | 8.65 | | | 182810 |
| RALGPS1 | NM_014636 | chr9 | 129677060 | 129985445 | 100 | 0.127 | 0.99986 | 0.96989 | 0.99081 | 18.81 | | | 614444 |
| ZER1 | NM_006336 | chr9 | 131492066 | 131534206 | 100 | 0.178 | 0.99877 | 0.96786 | 0.98184 | 26.16 | | | 617764 |
| SET | NM_003011 | chr9 | 131451500 | 131458675 | 100 | 0.183 | 0.96336 | 0.77029 | 0.99326 | 2.03 | | | 600960 |
| ZBTB43 | NM_014007 | chr9 | 129567318 | 129600487 | 100 | 0.225 | 0.87704 | 0.9283 | 0.78518 | 15.2 | | | 618676 |
| ENG | NM_001114753 | chr9 | 130577294 | 130616937 | 100 | 0.24 | 0.98962 | 0.8817 | 0.95072 | 23.71 | 3 (Sufficient Evidence) | 0 (No Evidence) | 131195 |
| DNM1 | NM_004408 | chr9 | 130965657 | 131017523 | 100 | 0.252 | 0.99986 | 0.97182 | 0.99969 | 13.49 | | | 602377 |
| GOLGA2 | NM_001366244 | chr9 | 131018107 | 131038231 | 100 | 0.295 | 0.99493 | 0.86155 | 0.76737 | 73.06 | | | 602580 |
| NUP188 | NM_015354 | chr9 | 131709977 | 131769375 | 100 | 0.297 | 0.98708 | 0.97359 | 0.98789 | 31.46 | | | 615587 |
| ZBTB34 | NM_001099270 | chr9 | 129622917 | 129648156 | 100 | 0.298 | 0.82962 | 0.91354 | 0.59235 | 20.93 | | | 611692 |
| LRRC8A | NM_019594 | chr9 | 131644411 | 131680317 | 100 | 0.347 | 0.9161 | 0.87388 | 0.98382 | 25.21 | | | 608360 |

Download gene table

Gene table ? ClinGen gene disease table ? ClinGen region table ? DECIPHER CNV syndromes ?

| Gene | Disease | Disease ID (MONDO) | Mode of inheritance | Classification | Date of classification | Online report |
|---|---|---|---|---|---|---|
| DNM1 | developmental and epileptic encephalopathy | MONDO:0100062 | AD | Definitive | 2016-12-13 | link |
| ENG | hereditary hemorrhagic telangiectasia | MONDO:0019180 | AD | Definitive | 2019-08-28 | link |
| LMX1B | nail-patella syndrome | MONDO:0008061 | AD | Definitive | 2021-07-27 | link |
| SPTAN1 | developmental and epileptic encephalopathy | MONDO:0100062 | AD | Definitive | 2019-01-17 | link |
| STXBP1 | developmental and epileptic encephalopathy | MONDO:0100062 | AD | Definitive | 2017-10-20 | link |
| ENG | generalized juvenile polyposis/juvenile polyposis coli | MONDO:0008276 | AD | Limited | 2017-12-11 | link |

**Fig. 5: Gene table and ClinGen gene-disease association table.** The gene table includes all intersecting genes, annotated gene scores, and links to their OMIM website. In the ClinGen gene-disease association table, all genes associated with a disease in ClinGen, their disease ID (MONDO), their mode of inheritance, and link to the ClinGen online report, are displayed.

Zooming back to the larger region, we filtered the CNVs based on their phenotypes. We identified that patients with small microdeletions that only intersect *STXBP1* to have epilepsy and intellectual disability. At the same time, finger dysplasia and facial dysmorphism could only be seen in patients with larger CNVs, indicating more driver genes involved in their phenotypes (Fig. 6). The CNV-ClinViewer showed more genes in this region are associated with autosomal dominant diseases (*DNM1, ENG, SPTAN1* and *LMX1B*) (Fig. 5) which aligns with the conception that 9q34.11 genomic deletions involving *ENG*, *TOR1A*, *STXBP1*, and *SPTAN1* are revealing cis-genetic effects leading to complex phenotypes[1].

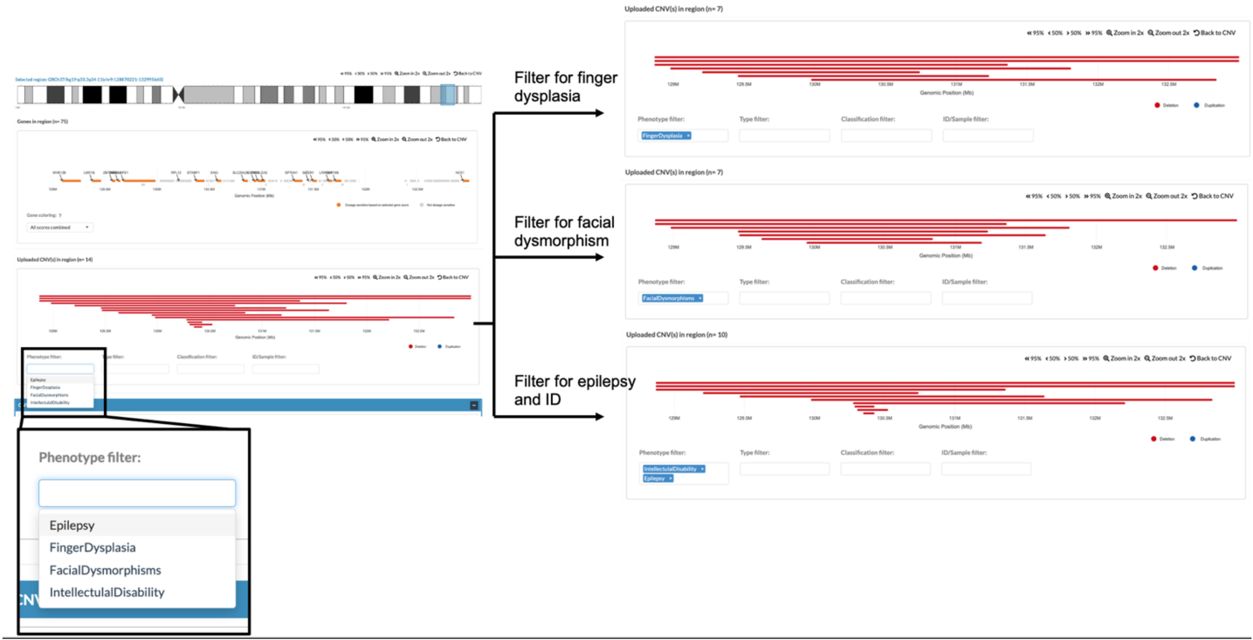


**Fig. 6: Dynamic filtering based on phenotypes of uploaded and visualized CNVs.** If binary phenotype information is provided in the input data, users can filter the CNVs based on those to perform genotype-phenotype analyses.

Besides the evaluation of the gene content, we could use the CNV-ClinViewer to identify overlap with 55 pathogenic/ likely pathogenic CNVs in ClinVar (15 deletions with 100% in the region), explore their phenotypes and get directed to the ClinVar variant websites for more information (Fig. 7).

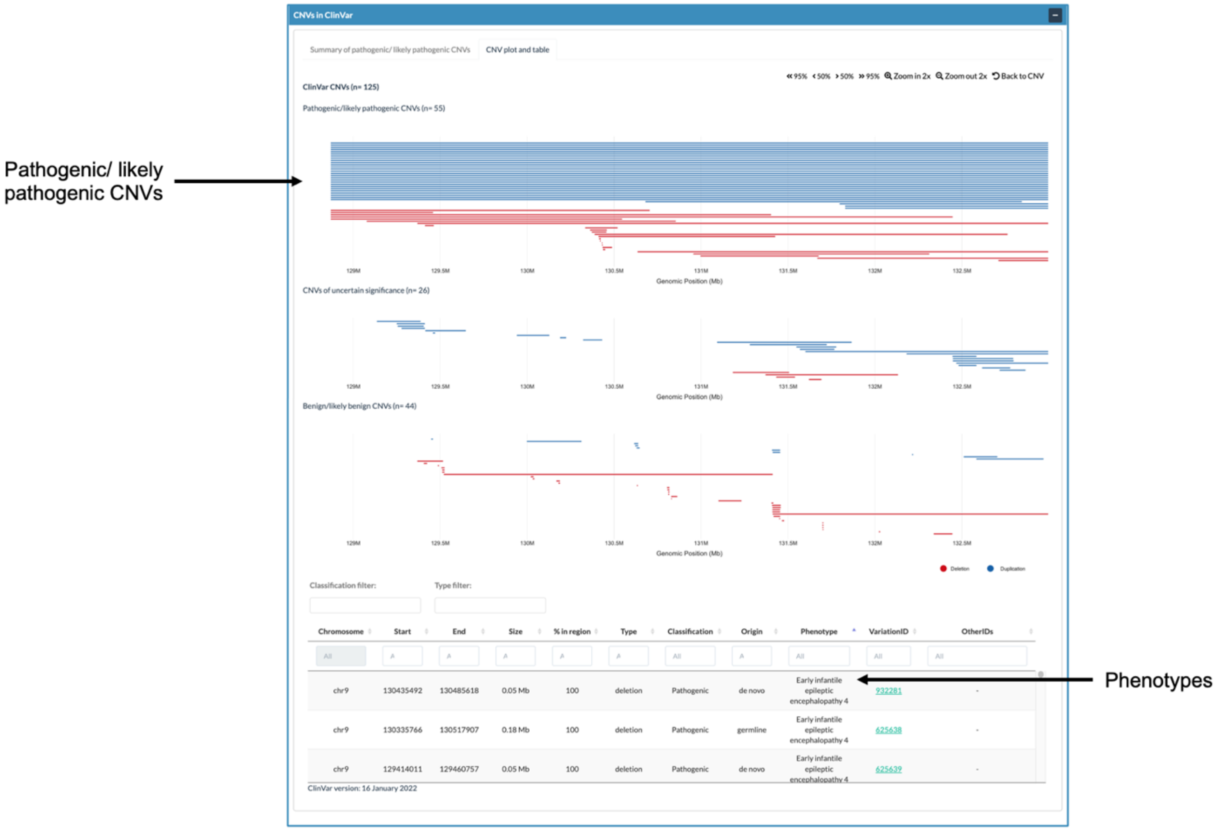


**Fig. 7: Identification of overlap with ClinVar CNVs**. The ClinVar track in the genomic viewer shows all intersecting CNVs from ClinVar, grouped by their clinical significance. The ClinVar CNVs can be filtered based on their classification and type. Detailed information about the phenotypes, origin, and links to their ClinVar reports can be found in a table below the visualization.

Overall, the CNV-ClinViewer successfully assisted the interpretation, exploration and analysis of the uploaded CNVs, and could replicate research findings that otherwise would require the usage of different databases and tools.
